# Agreement on Respiratory Impedance Measured by Two Commercial Forced Oscillation Technique Devices: A Randomized Crossover Study

**DOI:** 10.1101/2025.04.03.25325095

**Authors:** Sajal De

## Abstract

**Background:** The performance of forced oscillation technique (FOT) devices may vary due to differences in hardware and signal processing.

**Research question:** Does the respiratory impedance (Zrs) measured by two commercial FOT devices agree?

**Study Design and Methods:** The Zrs of 97 consecutive adults with respiratory symptoms were measured using TremoFlo^®^ C-100 and Resmon™ Pro FULL V3, employing a random allocation and crossover design. The parameters between the devices were compared using paired t-tests and Bland-Altman plots with 95% limits of agreement (LoA).

**Results:** The average age of the study population was 42.3 years, with 46% were women. The resistance parameters (R5 and R19) of Resmon^™^ were larger than those obtained with TremoFlo^®^. However, R5-19 of Resmon^™^ was significantly smaller than TremoFlo^®^, with a paired difference of -0.20 cm H_2_O/L/s (95% CI: -0.35, -0.05; p = 0.01). The within- and whole-breath X5 of Resmon^™^ were significantly less negative than those obtained with TremoFlo^®^. The reactance area and resonant frequency of Resmon^™^ were also significantly smaller than those obtained with TremoFlo^®^. The bias for R5 (Bias: 0.29 cmH_2_O/L/s; LoA: - 2.69, 3.26) was broad and had random errors. The bias for X5 (bias: 0.8 cmH_2_O/L/s; 95% LoA: -1.25, 2.85) and Ax (Bias: -13.66 cmH_2_O/L/s; LoA: -45.46, 18.14) was also wide and exhibited systematic errors.

**Interpretations:** Respiratory impedance measured by the two FOT devices differed significantly, particularly for the reactance parameters. Normative Zrs generated by a particular oscillometer should be avoided when interpreting reports obtained from another device.

## Background

Lung oscillometry is becoming popular for the non-invasive assessment of lung function, particularly in patients with obstructive airway diseases. The oscillometer measures the impedance of the respiratory system (Zrs) by superimposing pressure or sound waves of multiple frequencies on tidal breaths. The Zrs comprise respiratory system resistance (Rrs) and respiratory system reactance (Xrs).^1^ The Impulse Oscillometry System (IOS) utilizes patented rectangular waves, distinguishing it from other oscillometers that employ sinusoidal waves, such as forced oscillation technique (FOT) devices.

Performance across oscillometers may vary due to differences in hardware and signal processing software.^1^ The European Respiratory Society (ERS) guidelines recommend extensive validation of the oscillometer in various conditions, including healthy and diseased states.^1^ The normative Zrs generated by a particular oscillometer may not be useful for interpreting the results obtained from another oscillometer.^1^ Previous studies have mostly assessed the agreement between Zrs measured by the TremoFlo^®^ C-100, a FOT device, and the IOS.^2–4^ Only one study has evaluated the agreement of Zrs measured by two FOT devices in a small number of patients.^5^. It is unclear whether those findings can be generalized to patients with respiratory symptoms who typically undergo lung oscillometry for diagnostic workups.

This study examined the agreement between respiratory impedance measurements using TremoFlo^®^ C-100 (THORASYS Thoracic Medical Systems Inc.) and Resmon^™^ Pro FULL V3 (Restech Srl, Milan, Italy) in adults with respiratory symptoms. The hypothesis was that since both were FOT devices, the Zrs measured by them should differ little, and the bias (if any) should be random.

### Study Design and Methods

This prospective single-center, randomized crossover study was conducted from January 2025 to February 2025. The Institutional Ethics Committee of All India Institute of Medical Sciences, Raipur, India (4414/IEC-AIIMSRPS/2024) approved the study protocol. This study was conducted in accordance with the Declaration of Helsinki.

### Study participants

Consecutive adults aged >18 years with respiratory symptoms (chronic cough, breathlessness, etc.), bronchial asthma, and chronic obstructive pulmonary disease (COPD) were recruited from the outpatient department. Written informed consent was obtained from each participant.

### Sample size calculation

The study required 97 patients to achieve a power of 90% and a significance level of 5% (two-sided) to detect a paired mean difference of 0.2 cmH_2_O/L/s, assuming the standard deviation of the differences of 0.6 cmH_2_O/L/s.^6^

### Data Collection

Anthropometric data (age, gender, height, and weight), clinical diagnosis, or indication for oscillometry were recorded. Patients were randomly assigned to an AB or BA sequence using Random Allocation Software 2.0 (https://random-allocation-software.software.informer.com/2.0/). Patients in the AB sequence were studied first with Resmon^™^ Pro, followed by TremoFlo^®^, and vice versa in the BA sequence. Both devices were calibrated according to the manufacturer’s instructions before use. Oscillometry was performed in an upright sitting position, with checks supported by the hands, wearing a nose clip, and in accordance with the ERS guidelines.^1^ Clearflo^™^ F-100 antibacterial filters were used for both devices. Each patient performed at least three sessions for each device. The Resmon version 21.5.0 and Tremolo version 1.2 software were used to analyze the Zrs.

### Parameters

The parameters examined in this study were total Rrs (R5) and Xrs (X5) at 5 Hz, inspiratory and expiratory Xrs at 5 Hz, the difference between inspiratory and expiratory Xrs at 5 Hz (ΔX5), the difference in Rrs at 5 Hz and 19 Hz (R5-19), the reactance area (Ax), resonant frequencies (Fres), and coefficient of variations (CoV) for R5. The current TremoFlo^®^ software does not allow for the independent calculation of ΔX5. Therefore, the expiratory X5 value was subtracted from the inspiratory X5 value to calculate the ΔX5 for TremoFlo^®.^

Oscillometry was repeated 15–20 min after inhaling 400 μg of salbutamol using a metered-dose inhaler through a spacer device to assess the bronchodilator responsiveness (BDR) in patients with suspected obstructive airway diseases. BDR was defined according to the ERS technical standard, i.e., a 40% decrease in R5, a 50% decrease in X5, and an 80% decrease in Ax as per the ERS standard.^1^

### Statistical Analysis

The data were analyzed using MedCalc^®^ statistical software for Windows, version 23.0.8 (MedCalc Software, Ostend, Belgium) and GraphPad Prism 8 (GraphPad Software, Inc., La Jolla, CA, USA). The data were expressed as the mean ± standard deviation and number with percentage. The difference in Zrs was compared using paired t-tests with a 95% confidence interval (CI). Bland-Altman analysis was employed to visualize the agreement and bias between the two devices. The upper and lower limits of agreement (LoA) were defined by the 95% confidence interval (CI) of the differences, calculated as the mean difference ± 1.96 standard deviations of the difference. The acceptable limits of agreement for R5 and X5 were considered to be 0.2 cmH_2_O/L/s. The consistency between the BDR by both devices was evaluated using Cohen’s kappa (κ). The values of κ 0.41– 0.60, 0.61–0.80, and 0.81-1.0 indicate moderate, substantial, and perfect agreement, respectively. A p-value <0.5 was considered statistically significant.

## Results

### Patients Characteristics

We recruited 113 consecutive adult patients. Among them, 97 patients had a CoV of R5 < 10% for both devices and were included in this study. Sixty patients (62%) were evaluated for their respiratory symptoms; 31% had a clinical diagnosis of bronchial asthma, and 7% had COPD. The mean age of the study population was 42.3 ± 15.5 years, with 46% were female. The body mass index of the cohort was 24.7 ± 4.1 kg/m².

The mean CoV of R5 for Resmon™ and TremoFlo^®^ was 6.5% and 5.7%, respectively. The paired t-test of Zrs between the devices is shown in Figure 1. The R5 of Resmon™ was not significantly higher than that of TremoFlo^®^, with a paired difference of 0.29 cmH_2_O/L/s (95% CI: - 0.02, 0.59; p = 0.07). The R19 of Resmon^™^ was significantly higher than that of TremoFlo^®^, with a paired difference of 0.48 cmH_2_O/L/s (95% CI: 0.28, 0.68; p<0.001). The R5-19 of Resmon^™^ was significantly smaller than TremoFlo^®^ with a paired difference of -0.20 cmH_2_O/L/s (95% CI: -0.35, -0.05; p = 0.01).

**Figure 1.**
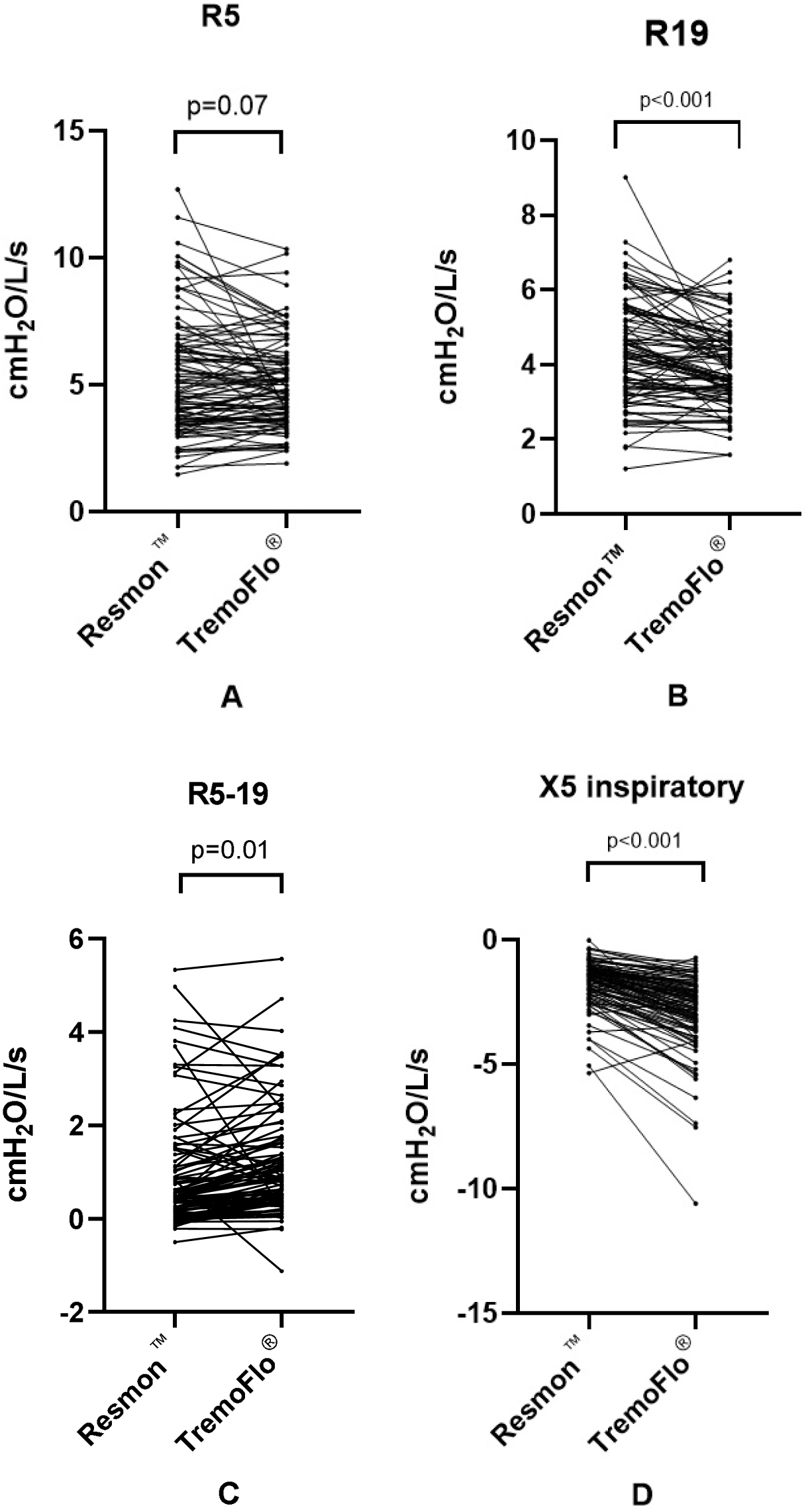
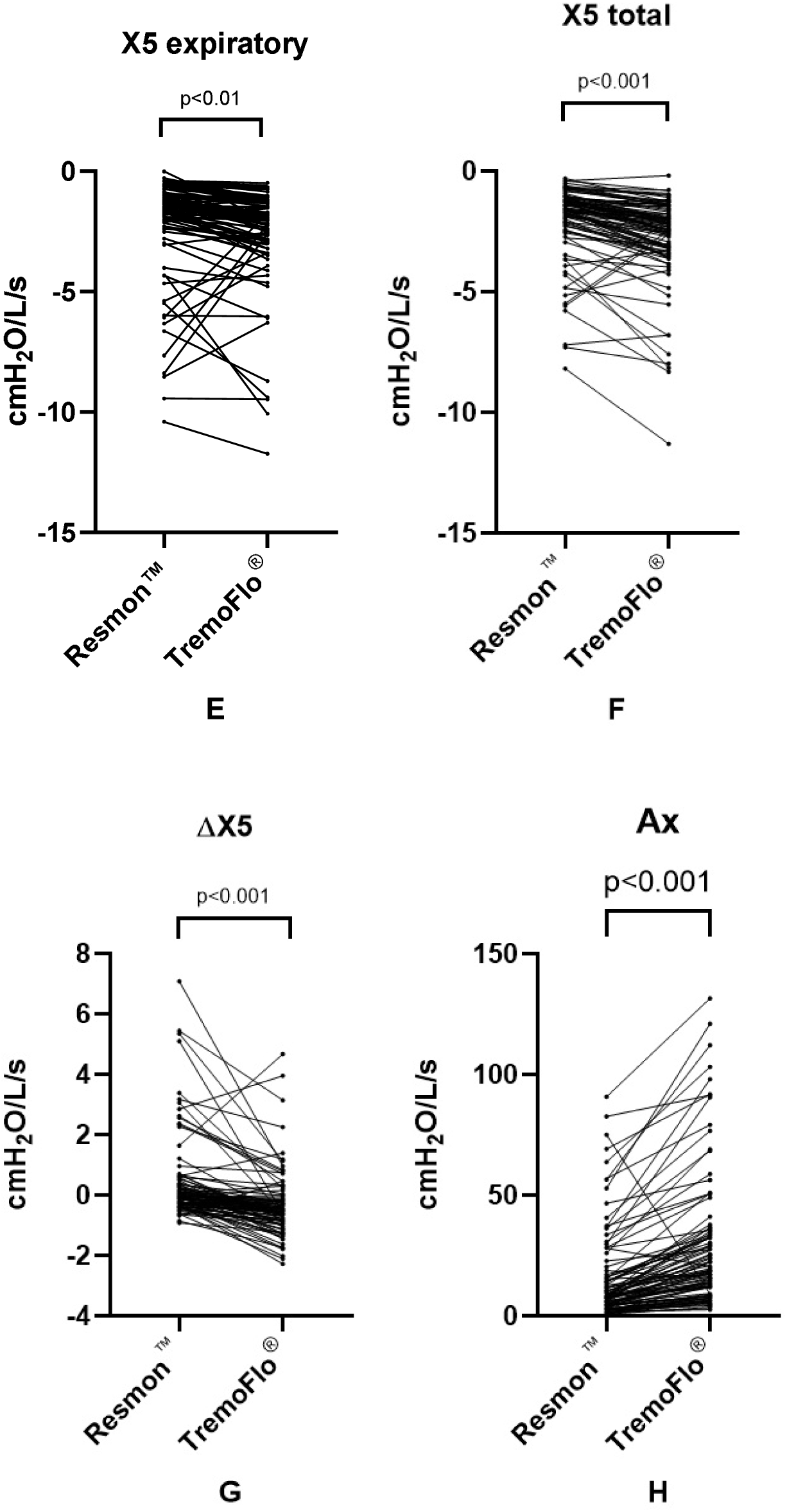
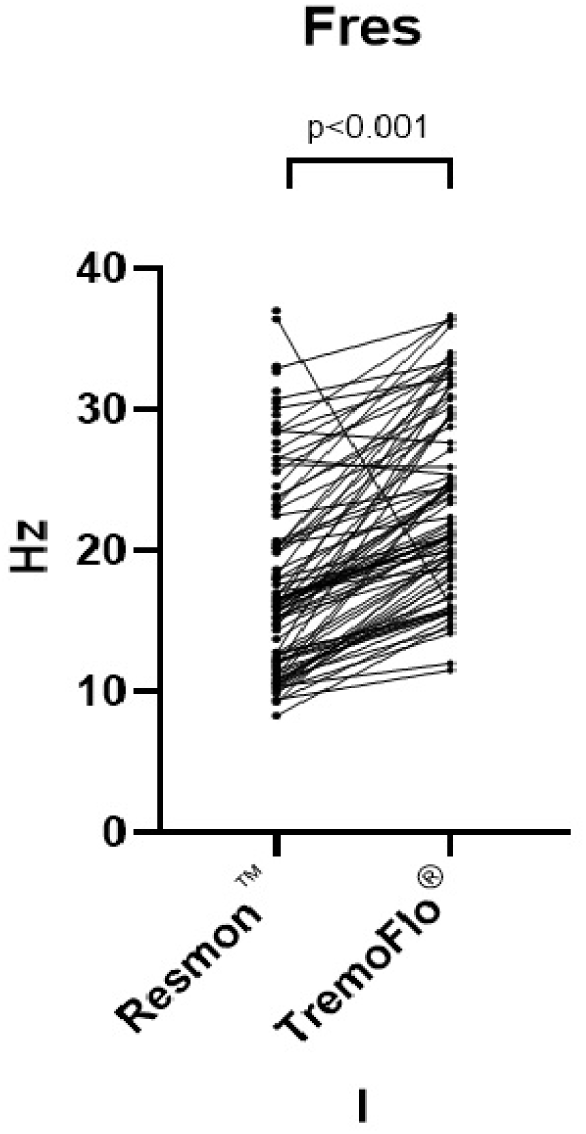
Paired t-test of respiratory impedance parameters measured by Resmon^™^ and TremoFlo^®^. Each pair of points connected by a line represents one patient. (A) R5; (B) R19; (C) R5-19; (D) X5inspiratory; (E) X5expiratory; (F) X5 total; (G) ΔX5; (H) Ax; and (I) Fres.

The within- and whole-breath X5 of Resmon™ were significantly less negative than those obtained from TremoFlo^®^. The paired differences between Resmon™ and TremoFlo^®^ of X5 inspiratory (1.26 cmH_2_O/L/s [95% CI: 1.06-1.46, p<0.001]), X5 expiratory (0.46 cmH_2_O/L/s [95% CI: 0.16, 0.76; p<0.01]), and X5 total (0.8 cmH_2_O/L/s [95% CI: 0.59, 1.01; p<0.001]) were large. The large differences in Xrs between the devices were clinically relevant. The ΔX5 of Resmon™ was significantly larger, with a paired difference of 0.83 cmH_2_O/L/s (95% CI: 0.59, 1.07; p <0.001).

The Ax and Fres of Resmon™ were significantly smaller than TremoFlo^®^ with paired differences of -13.66 cmH_2_O/L/s (95% CI: -16.93, -10.39; p<0.001) and - 6.51 Hz (95% CI: - 7.62, -5.39; p<0.001), respectively.

The Bland-Altman plots of agreement between Zrs from the devices are shown in Figure 2. The Bland-Altman plot revealed that the bias of R5 between the devices was wide (0.29 cmH2O/L/s; 95% LoA: -2.69, 3.26) and exhibited random errors. The bias of R19 was also wide and exhibited random errors (bias: 0.48 cmH_2_O/L/s; 95% LoA: -1.65, 1.25). The R5-19 between the devices showed a systematic bias of -0.2 cmH_2_O/L/s (95% LoA: -1.51, 2.47). The bias of X5 total (bias: 0.8 cmH_2_O/L/s; 95% LoA: -1.25, 2.85) was evident with consistent and systematic errors, particularly at higher values. The bias of X5 inspiratory (bias: 1.26 cmH_2_O/L/s; 95% LoA: -0.66, 3.18), X5 expiratory (bias: 0.46 cmH_2_O/L/s; 95% LoA: -2.41, 3.33), and Δ X5 (bias: 0.83 cmH_2_O/L/s; 95% LoA: -1.5, 3.16) also wide and had systematic errors. The bias of AX (-13.66 cmH_2_O/L/s; 95% LoA: -45.46, 18.14) and Fres (- 6.51 Hz; 95% LoA: -16.63, 3.62) was wide, with consistent systematic errors. The LoA of Xrs was much broader at higher values.

**Figure 2.**
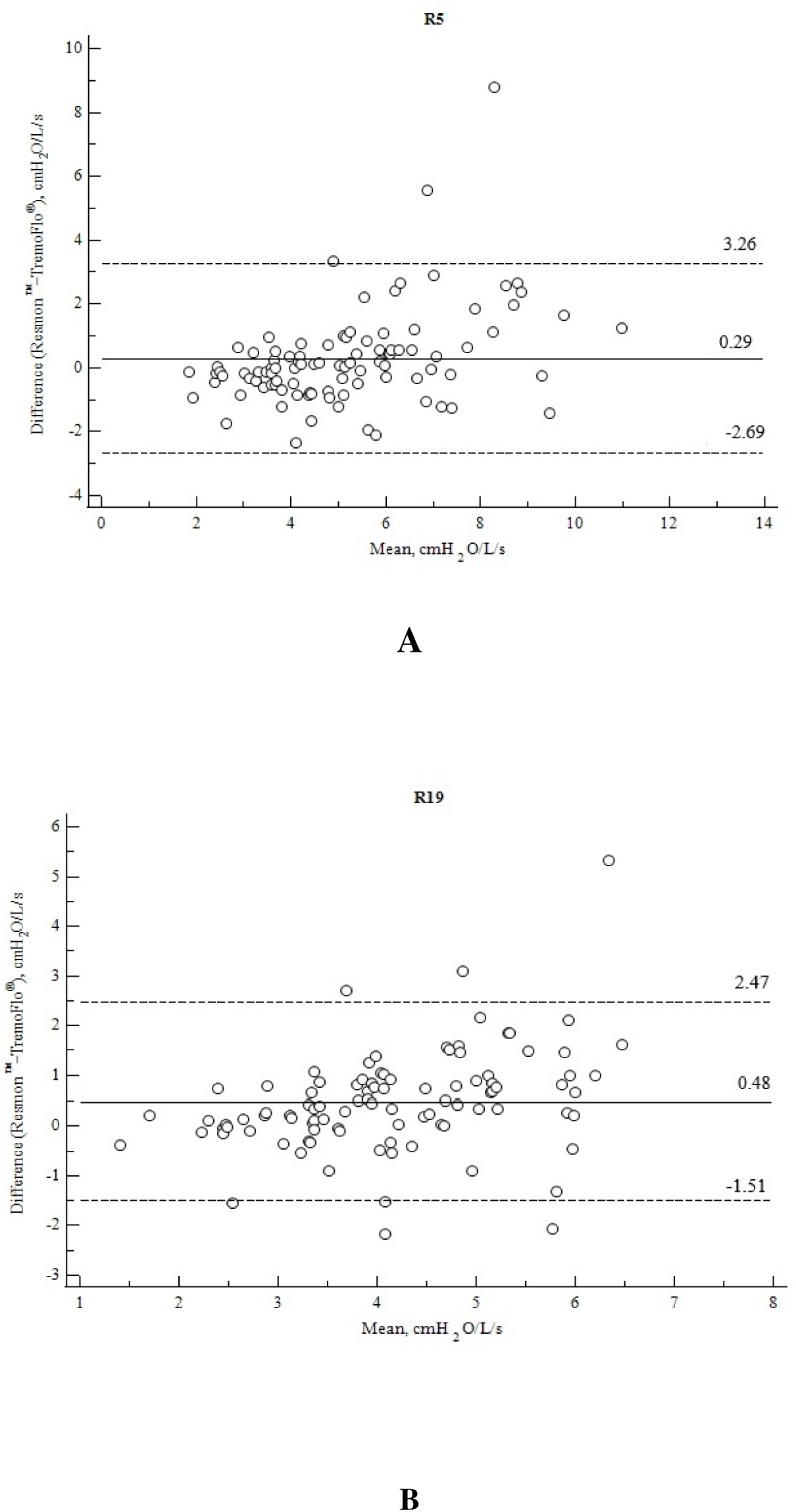
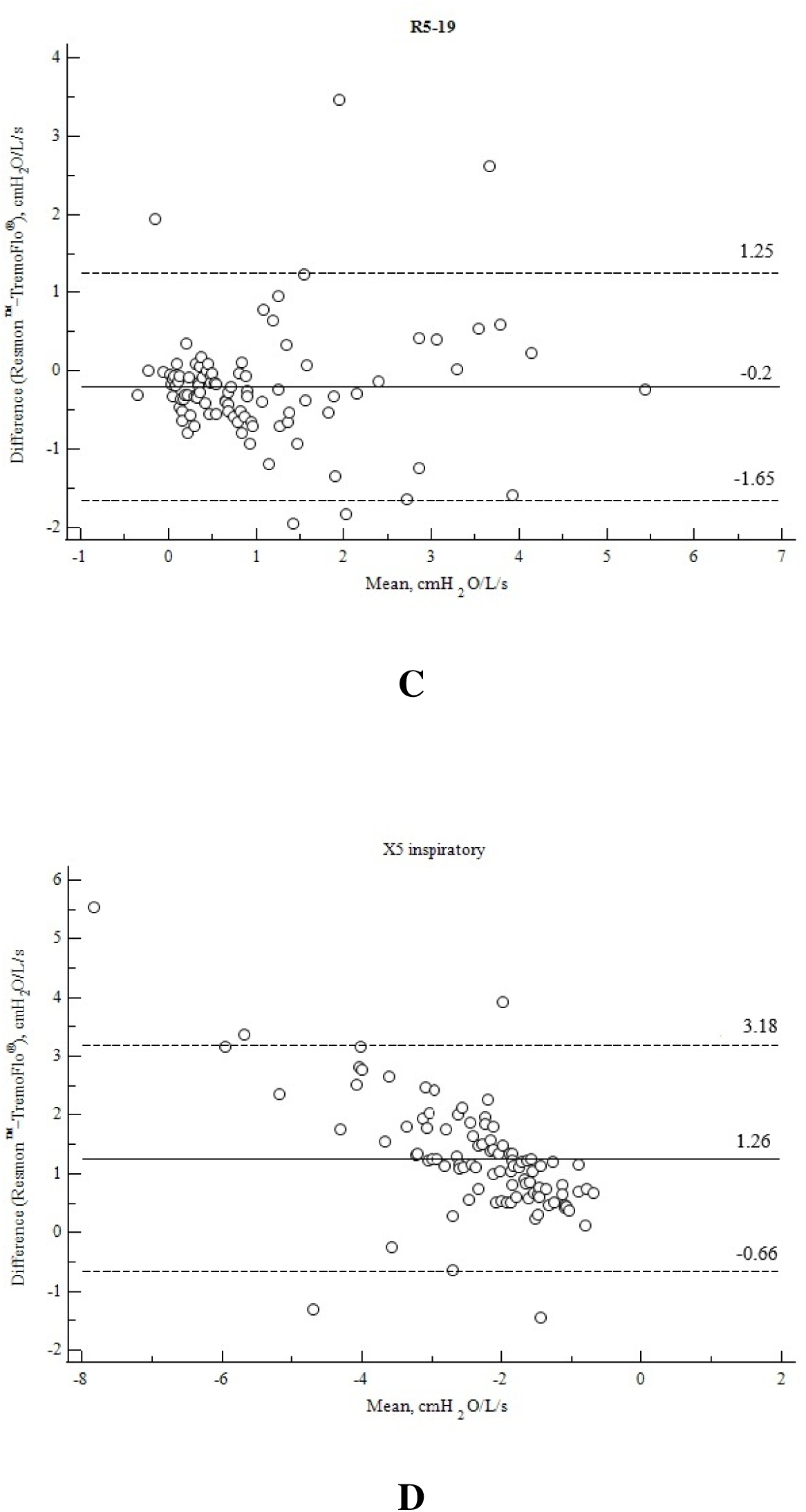
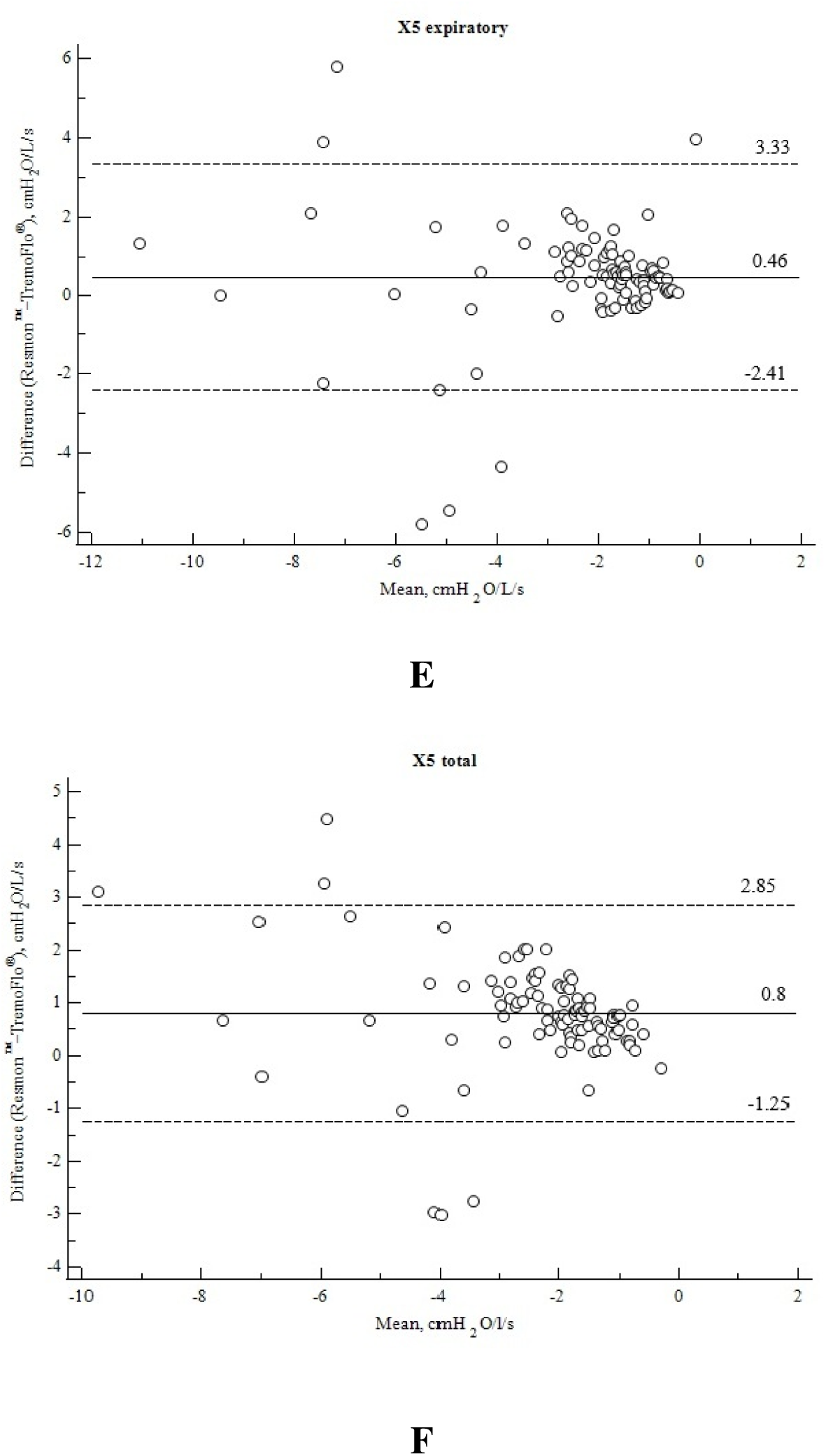
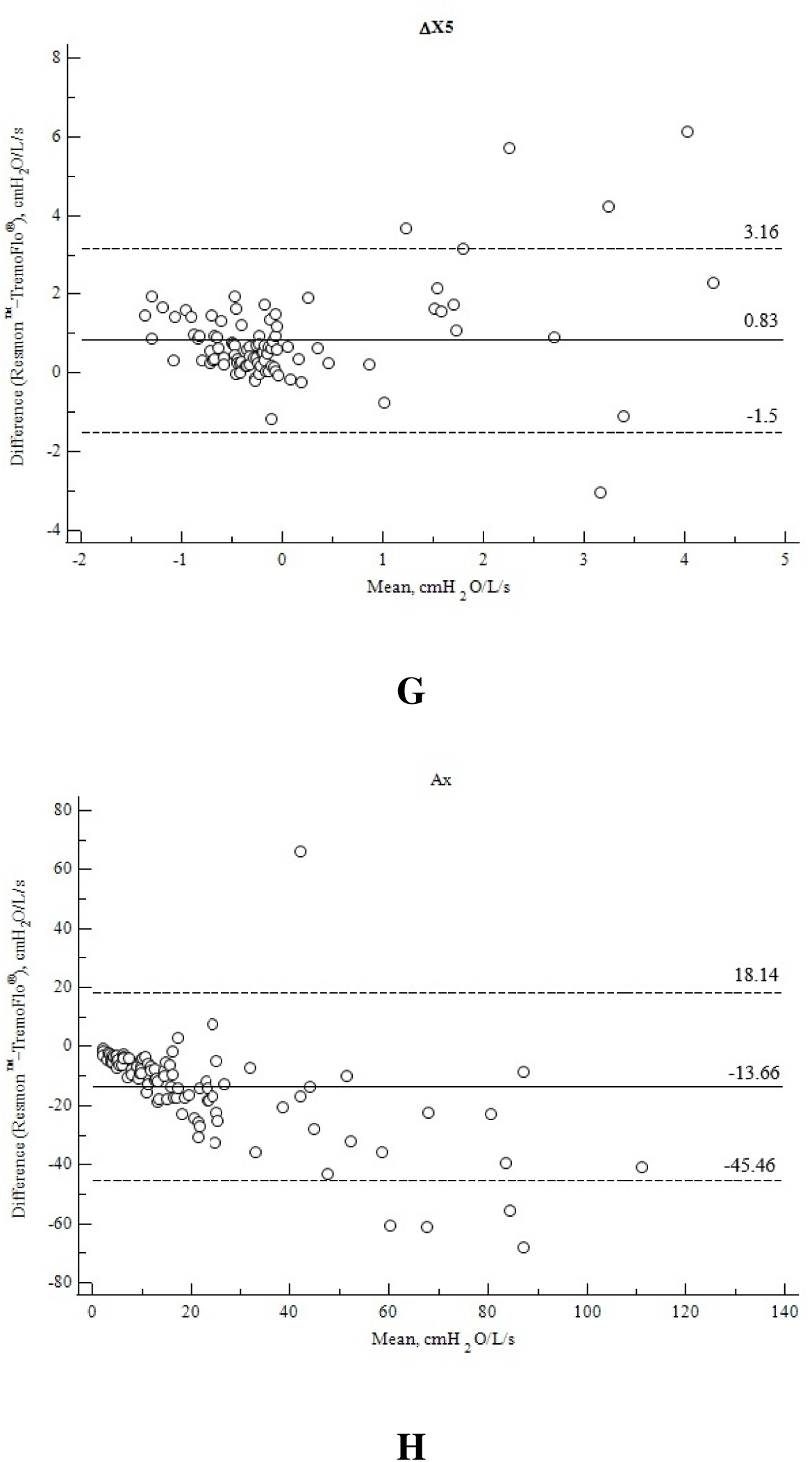
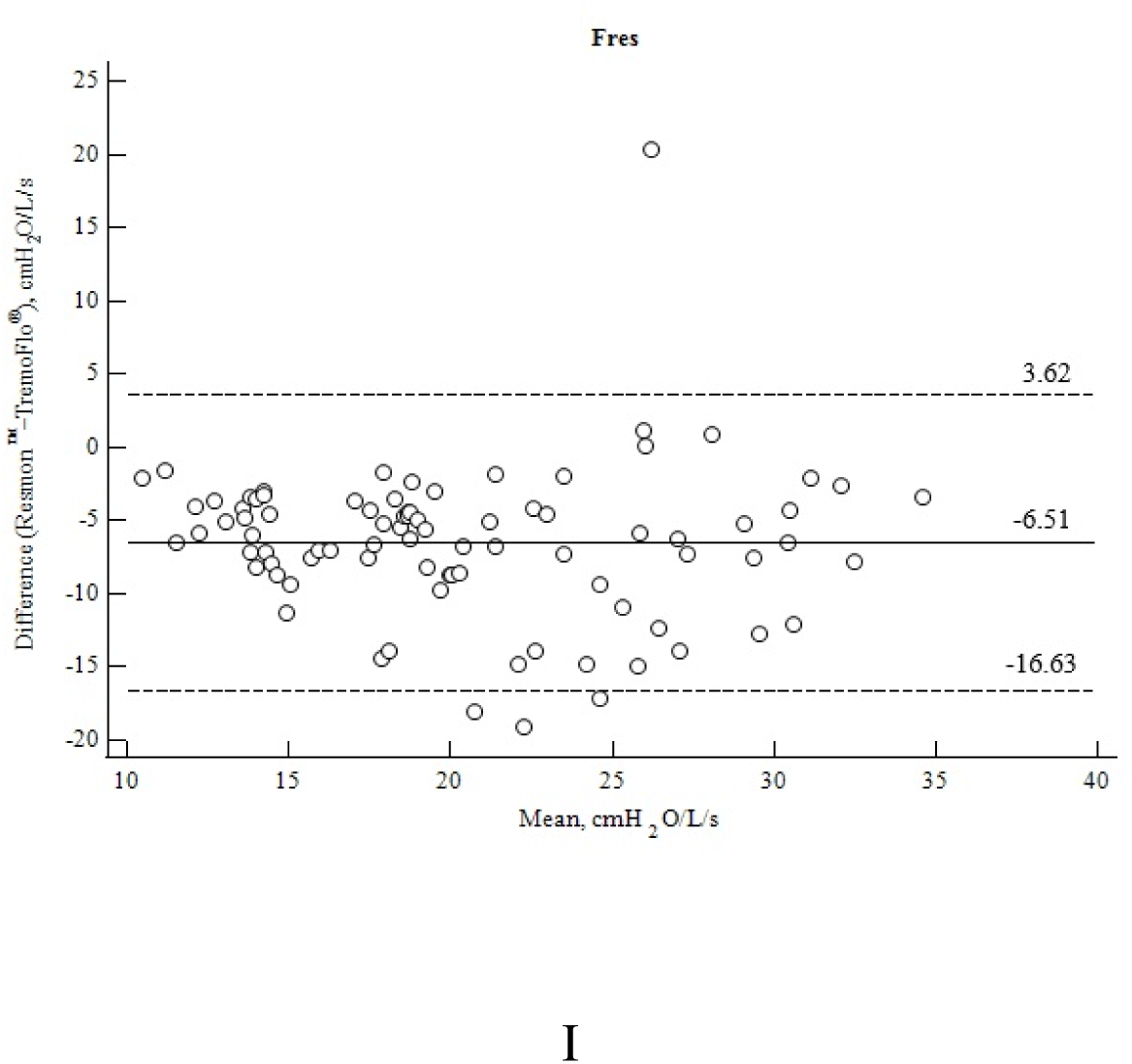
Bland-Altman plots with 95% limits of agreement of respiratory impedance measured by Resmon^™^ and TremoFlo^®^ (A) R5; (B) R19; (C) R5-19; (D) X5 inspiratory; (E) X5 expiratory; (F) X5 total; (G) ΔX5; (H) Ax; and (I) Fres. Solid lines represent the mean difference between measurements, and dashed lines represent the 95% limits of agreement.

BDR was performed in 58 patients. The proportion of BDR of R5 in Resmon™ was significantly higher than that in TremoFlo^®^ (20.7% vs. 17.2%, p = 0.003), and the agreement was moderate (κ = 0.44). The proportion of BDR for both X5 and AX between the devices was equal (7%) and showed a perfect agreement (κ = 1.0).

## Discussion

This study investigated the agreement between the Zrs obtained from TremoFlo^®^ C- 100 and Resmon^™^ Pro FULL V3 in a clinical setting. This study revealed that although both are FOT devices, there was considerable variability in Zrs. The Rrs, especially R19 of Resmon™, were significantly higher. The bias in measuring R5 between the devices was random. The Resmon™ recorded significantly less negative Xrs (X5 inspiratory, X5 expiratory, X5 total), smaller Ax, and smaller Fres than TremoFlo^®^. The LoA of Xrs was wide and showed a systematic bias. The wide LoA indicates that measurements of Zrs were device-specific.

Previous studies mostly examined the agreement between TremoFlo^®^ and the IOS.^2–4,7^ Ducharme et al. examined the agreement between TremoFlo^®^ and the IOS in asthmatic children.^2^ TremoFlo^®^ recorded significantly higher R19, more negative X5, and smaller R5- 20, with a bias of ≥30%. The paired difference of R5 in their study was nil but had a proportional bias. TremoFlo^®^ recorded a relatively more negative X5, larger Fres and AX, and smaller R5 compared to the IOS in a study by Kuo et al.^3^ Soares et al. compared TremoFlo^®^ with the IOS among healthy adults, asymptomatic smokers, individuals with bronchial asthma, and a 3D airway model.^4^ TremoFlo^®^ recorded smaller R5, R20, and R5-20 than the IOS. The differences were small and exhibited significant systematic bias. The X5 of TremoFlo^®^ was significantly more negative and had a larger AX than the IOS. Lundblad et al. observed that R5 and R5-19 of TremoFlo^®^ were higher for COPD but smaller in healthy than the IOS.^7^ The X5 of TremoFlo^®^ was significantly more negative in COPD patients but less negative in healthy individuals. Overall, X5 of TremoFlo^®^ was more negative, Ax and Fres were higher than the IOS. Previous studies mostly documented that the X5 of TremoFlo^®^ was more negative than the IOS, while the agreement of R5 with the IOS across the studies was inconsistent.

Zimmermann et al. assessed the agreement of Resmon^™^, TremoFlo^®^, and the IOS with a custom-built device in both in vivo and in vitro settings.^5^ Contrary to the current study, the R5 of Resmon^™^ in their study was smaller than that of TremoFlo^®^, but the difference was statistically insignificant. Similar to the present study, the X5 total, X5 inspiratory, and X5 expiratory values of TremoFlo^®^ were significantly more negative than those of Resmon^™^. The X5 inspiratory of TremoFlo^®^ in the current study was more negative than the X5 expiratory. As a result, expiratory flow limitation at tidal breaths, i.e., ΔX5 > 0.28 kPa/L/s, was detected in a smaller number of cases by TremoFlo (3.2%) compared to Resmon (7.2%). Zimmermann et al. also observed that ΔX5 of TremoFlo^®^ was more negative than that of the IOS.^5^

Dandurand et al. observed that as test loads increase, the difference in Zrs measurements across the oscillometer increases.^8^ The authors proposed that unknown instrumental and signal processing factors may be responsible for the significant differences between the devices. Similar to earlier studies, the present study also demonstrated more variability between devices in patients with high Zrs.^3,4,8^ The paired difference between the parameters of this study was higher than in earlier studies as the study population was heterogeneous.

The Xrs and R5-19 were used to assess the small airway dysfunction. Both were significantly higher in TremoFlo^®^ than those of Resmon^™^, especially at higher values. Therefore, there is a risk of overestimating small airway dysfunction by TremoFlo^®^ among patients with obstructive airway diseases. The homogeneity of the study population in previous studies may limit the relevance of those studies for real-life diagnostic applications of oscillometers. Since the earlier studies were published, both devices have undergone software upgrades.

The differences in measurements across devices can be attributed to variations in breathing cycles, CoV, signal processing, and data analysis [1, 5, 8]. The CoV of R5 for both devices was <10%. Therefore, the observed differences between the devices in the present study were likely due to variations in the data collection and algorithms used for analysis. The different ethnicities of the study population may have contributed to the inconsistency in agreement across earlier studies.

This study arbitrarily defined the minimal clinically important differences (MCID) between the devices. The MCID for Zrs, particularly in healthy populations, is not defined. The threshold for MCID can be determined by the ratio of the difference divided by the standard deviation of a baseline score. A ratio of <0.2 is considered no change.^9^ Abdo et al. proposed a change in R5-20 ≥ 0.61 cmH_2_O/L/s and AX ≥ 6.6 cmH_2_O/L/s as MCID in patients with bronchial asthma.^10^ The bias of R5-19 in the current study was lower than the proposed MCID. This may be because the current study cohort was heterogeneous, with bronchial asthma patients comprising only one-third of the cohort. The bias in Ax of the current study was significantly higher than the proposed MCID.

The strength of this study lies in the inclusion of unselected patients with respiratory symptoms, the random order of measurements, and a CoV of R5 <10%. The random crossover design employed in the present study minimized the risk of internal inconsistencies. The antibacterial filters can affect the accuracy of the measurements. The antibacterial filter recommended by TremoFlo^®^ was used for both devices. Resmon™ measured the Zrs of the filter before each session, and the impedance of this filter ranged from 0.48 to 0.51 cmH_2_O/L/s. This study was the first to compare the agreement on BDR between the two devices.

### Interpretations

This study revealed both random and systematic biases in Zrs between the TremoFlo^®^ C-100 and Resmon^™^ Pro FULL V3. TremoFlo^®^ consistently and significantly overestimates the reactance parameters compared to Resmon^™^. The broader bias of reactance at higher values suggests that the agreement between the devices is less in patients with abnormal Zrs. The standardization of FOT devices using different test loads is required. Until then, normative values specific to the device should be used for clinical interpretations. The software upgrade may affect the measurements, which must be evaluated and reported accordingly.

## Data Availability

The author has no objection to sharing the data, provided it is done under confidentiality and with appropriate data protection and privacy, including with the journal statisticians upon request to verify or validate the analysis.

## Abbreviations

BDR: Bronchodilator responsiveness
COPD: Chronic obstructive pulmonary disease
MCID: Minimal clinically important differences
R5: Respiratory system resistance at 5 Hz
R5-19: Difference in respiratory resistance between 5 Hz and 19 Hz
Rrs: Respiratory system resistance
X5: respiratory system reactance at 5 Hz
Xrs: Respiratory system reactance
Zrs: Respiratory impedance
ΔX5: Difference between inspiratory and expiratory reactance at 5 Hz

